# World War II cohorts and diabetes mellitus, coronary heart disease and cerebrovascular disease later in life: An observational cohort study based on German claims data

**DOI:** 10.1101/2020.11.11.20227660

**Authors:** Thomas Fritze, Constantin Reinke, Gerard J. van den Berg, Gabriele Doblhammer

**Affiliations:** German Center for Neurodegenerative Diseases, Bonn, Germany; University of Rostock, Rostock, Germany; University of Groningen, Groningen, the Netherlands; University Medical Center Groningen, Groningen, the Netherlands; IFAU Uppsala, Uppsala, Sweden

## Abstract

**Background:** This study applied a data-driven approach to explore whether being born during or around World War II affects the risk of morbidity later in life.

**Methods:** Incident diagnoses were explored for diabetes (ICD-10 code E10-E14; 75,487 persons/12,905 incident cases), cerebrovascular disease (CeVD; I6; 79,829/11,664), and coronary heart disease (CHD; I20-I25; 89,657/11,116) for birth cohorts 1935-1950, using German health-claims data from 2004-2015. The data include quarterly information of the inpatient and outpatient treatment. We applied recursive partitioning methods with the aim of splitting the sample into birth cohorts with different risk profiles in terms of the incidence of diabetes, CeVD, and CHD. We performed Cox proportional hazard models to explore the transition into diabetes, CeVD, and CHD, accounting for birth cohort and gender. We used the median cut-points from the recursive partitioning analysis on the birth cohort indicator to define linear splines and present the yearly slopes of the linear splines.

**Results:** Based on the results from recursive partitioning method we defined four groups of birth cohorts depending on the disease of interest (diabetes: 1/35-10/37, 11/37-11/41, 12/41-10/44, 11/44-12/50; CHD: 1/35-5/37, 6/37-4/41, 10/44-12/50; CeVD: 1/35-6/37, 7/37-6/40, 7/40-12/43, 1/44-12/50). We found a general decline in cohort incidence for all three diseases, however, there was a discontinuation for diabetes among birth cohorts 12/41-10/44 (yearly slope=-0.01, p=0.584), and a deceleration for cohorts 11/44-12/50 for CeVD.

**Conclusions:** We identified risk periods associated with WWII that interfered with the cohort decline in the risk of diabetes and CeVD, stressing the importance of a beneficial early-life environment.

**Availability of data:** The scientific research institute of the AOK (WIdO) has strict rules regarding data sharing because of the fact that health claims data are a sensible data source and have ethical restrictions imposed due to concerns regarding privacy. Anonymized data are available to all interested researchers upon request. Interested individuals or an institution who wish to request access to the health claims data of the AOK, please contact the WIdO (webpage: http://www.wido.de/, mail).

## Introduction

Understanding life-long determinants of health at old age is of central importance in aging societies. Risk factors in early-, mid-, and late-life directly or indirectly trigger pathological changes in the body and subsequently lead to poor health later in life^1^. One approach to identifying possible risk factors is to explore cohort effects in those morbidity outcomes which are caused by exogenous events, such as famines^2^, wars^3^, or epidemics^4,5^. This study applies a data-driven approach in order to identify possible periods of risk associated with the Second World War (WWII). We use German health claims data to explore whether being born during or around WWII affects the risk of morbidity later in life. More specifically, we focus on cohort effects in the late-life risk of diabetes mellitus, cerebrovascular disease (CeVD), and coronary heart disease (CHD). These are some of the most common vascular and metabolic diseases and major risk factors regarding mortality, disability, and poor quality of life with high costs for the health care system^6-9^. We refer to the time around WWII as a sensitive period, with long-term consequences on late life health^10^.

### Living conditions in Germany during WWII

In Germany, the economy and nutrition differed comparatively little from pre-war conditions up until the year 1942, due to the exploitation of occupied regions in terms of food supply und forced labour in Germany. By 1943 agricultural production had fallen by only 10% in relation to 1939 and large contingents of food were being transferred from the occupied regions^11^. While ration cards existed from 1939 onwards^12^, caloric intake per person was about 2800 kcal daily until the winter of 1943/44, and thus was only 10% under the 1939/40 values. The quality of nutrition did change considerably, with calories coming less from meat and fat and more from vegetarian sources such as potatoes and cabbage. From the spring of 1942 rations of high-quality food were shortened massively, while from the winter of 1943/44 the nutritional situation deteriorated gradually due to the liberation of the occupied regions by the Allied Forces. This also meant that the German forces had to be supplied with inner-German food rations, to the disadvantage of the civilian population. The loss of agricultural areas, the destruction of machinery that was necessary for food production, as well as high immigration of millions of refugees led to a food crisis in Germany between 1944 and 1948^11,13^. This extended period of malnutrition caused severe health problems such as dystrophy, impaired growth, and increased susceptibility to infections^14,15^. In 1945, German food rations dropped significantly, hence the average daily number of calories reached its lowest level in the months immediately after the war^13,14^.

Air raids on German cities started at the beginning of the war and continued throughout the conflict. Early air raids were focused primarily on infrastructure, such as oil and railway targets. As a consequence of the Area Bombing Directive from the British Government’s Air Ministry, housing and infrastructure were destroyed extensively by allied bombing from 1942 onwards. The targeted cities were not only large, industrialized cities such as Cologne, Berlin, Hamburg or Dresden. In addition to economic aspects, other considerations such as weather conditions, visibility of landmarks, or susceptibility of city centres to area bombing with incendiary bombs meant that smaller cities were also attacked^16,17^. People lost their homes and suffered from war-related traumata such as displacement, mandatory military service, or war captivity^18^. Historical data from this era are notoriously incomplete, absent or manipulated for propaganda reasons. Therefore, it is difficult to demarcate the various episodes in terms of early-life conditions. This is a marked contrast to famine studies based on e.g. the Dutch Hunger Winter which was a singular event with well-known start and end dates^2^. Our statistical learning approach is ideally suited to deal with a lack of external justification of such dates.

### Research strategy

Based on the analysis of German health claims data, we use algorithms from statistical learning to classify the birth cohorts between 1935 and 1950 into groups with homogenous patterns within and heterogeneous patterns between these groups regarding the risk of later-life diabetes, CeVD, and CHD. In the first instance, we perform recursive partitioning methods to distinguish cohorts according to their risk profiles. Then, taking the results from partitioning into account, we apply regression splines in Cox models to analyse the cohort effects on the relative risks of the diseases mentioned above. Given the general deterioration of the living circumstances from 1942 onwards, we expect a higher disease load in cohorts born between 1942 and the end of the war, as well as those born immediately after the war.

## Data and Methods

### Data

We analysed the incidence of diabetes, CeVD, and CHD using routine claims data from 2004-2015 of the largest German statutory health insurance, the “Allgemeine Ortskrankenkasse” (AOK). In Germany, 70 million people are covered through statutory health insurance policies (about 84.7% of the total population); about one-third of these people are insured through the AOK^19^.

A random sample of 250,000 insured persons born in or prior to 1954, and who had at least one day of insurance coverage by the AOK in the first quarter of 2004, was drawn by the WIdO (Scientific Institute of the AOK). Using a unique person ID, these individuals were followed quarterly over time between 2004 and 2015, establishing a longitudinal sample. The data include complete records of the inpatient and outpatient treatment that each insured patient received in this period.

### Outcome variables

We explored the incidence of three diseases that are defined according to the codes of the International Classification of Diseases, 10th Revision (ICD-10): diabetes mellitus (E10–E14); CeVD (I6); and CHD (I20-I25). We confirmed diagnoses by co-occurrence over time as an internal validation procedure in order to rule out false-positive diagnoses because we were not able to validate diagnoses by face-to-face examinations. Incident cases were defined as the first occurrence of a valid diagnosis. Furthermore, to avoid confusion between firstly-diagnosed incident cases and prevalent cases with a history of the respective disease, a period of at least two years without a valid diagnosis was warranted. Prevalent cases with a respective diagnosis in 2004 or 2005 were deleted from the sample. Thus, incident diagnosis of the respective disease is defined for all persons who had not had a valid diagnosis in 2004 and 2005, and who were diagnosed for the first time between 2006 and the second quarter of 2013 as the period of analysis. This ensures enough time to receive repeated diagnoses following the incident diagnosis.

### Exposure variable

We included a birth cohort indicator distinguishing month and year of birth. The indicator ranges from 1 to 192, equalling 1 for patients born in January 1935, 2 for patients born in February 1935, and so on, so that the value 192 is assigned to persons born in December 1950.

### Statistical analysis

In this longitudinal analysis, incident diagnoses were explored for the respective diseases for birth cohorts 1935-1950 at ages 55 and above. Analysis time calculated in years started at age 55 and ended at the time of the first diagnosis of the respective disease. In the case of no diagnosis, analysis time was censored at the age of death, the age of leaving the health insurance company, or the age at the end of the study period, June 30, 2013, whichever occurred first. As information on diagnoses was aggregated on a quarterly basis, the incidence of the respective disease was set in the middle of the respective quarter (which corresponded to 1.5 months in terms of analysis time). In the case of death, the time of death was assumed to be in the middle of the respective month (0.5 months in terms of analysis time).

Statistical analysis was then applied in three steps. First, to visually explore cohort trends in diabetes mellitus, CHD, and CeVD we calculated incidence rates by birth cohort. The Hodrick-Prescott Filter^20^ was applied with a smoothing value of 14,400 to separate the time series into trend and cyclical components. Since these trends mainly reflect the age dependency of morbidity (i.e. later-born cohorts are younger when we observe them in our data set), we expect a continuous decline in disease load from older to younger cohorts. Any deviation from this trend is then a cohort effect.

Second, to identify sensitive periods we applied recursive partitioning methods (survival trees) using the *ctree* function in the ‘partykit’-package (Version 1.5) in the statistical program R (Version 3.6.1). The package implements conditional inference trees^21,22^. We included only the birth cohort indicator, with the aim of splitting the sample into birth cohorts with different risk profiles in terms of the incidence of diabetes, CeVD, and CHD, respectively. The split criterion was based on log-rank statistics. Because we were interested in the most important splitting-values (sensitive periods) rather than predictions, we only consider the splitting values of the first three internal nodes. All parameters of *ctree* remained to default. To reduce the high variance by a single tree we used 1,000 bootstrapped samples with 10,000 cases per sample, averaging the splitting-values for each of the three nodes. Therefore, conditional inference trees use a concept of statistical significance. For the stopping criterion the number of resulting nodes may vary over the bootstrapped samples. We present the median as well as the 25^th^ and 75^th^ percentiles of the splitting values. Missing nodes are indicated by NA (‘not available’).

In the third step, we performed Cox proportional hazard models to explore the transition into diabetes, CeVD, and CHD respectively, and to calculate the risk of an incident diagnosis while accounting for birth cohort and gender. We controlled for left truncation by including the age in 2006 in the analysis time. We used the median cut-points from the recursive partitioning analysis on the birth cohort indicator to define linear splines. Using the results from Cox regression, we present the yearly slopes of the linear splines.

## Results

The number of explored subjects, person-time, and incident cases differed according to the respective disease. For diabetes, we had information on 75,487 patients with 466,789 person-years and 12,905 incident cases. The overall incidence rate was 27.65 cases per 1,000 persons (95% confidence interval (CI)=27.17 - 28.13). Exploring CHD as variable of interest, we had information on 79,829 patients with 498,272 person-years, 11,664 incident cases, and an overall incidence rate of 23.41 cases per 1,000 persons (95%-CI=22.99 - 23.84). We followed 89,657 patients over time and analysed 569,711 person-years and 11,116 incident cases of CeVD. The overall incident rate was 19.51 cases per 1,000 persons (95%-CI=19.15 - 19.88) (Table 1).

**Table 1:**
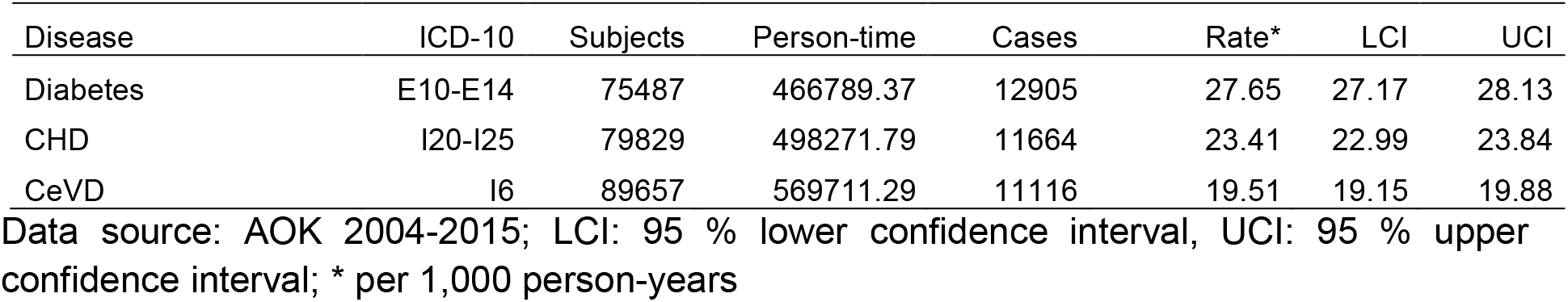
Number of subjects and cases, person-time and incidence rates

The overall morbidity pattern revealed an almost linear decline in the disease-specific incidence rates over birth cohorts, with some small deviations in the period of WWII (Figure 1).

**Figure 1:**
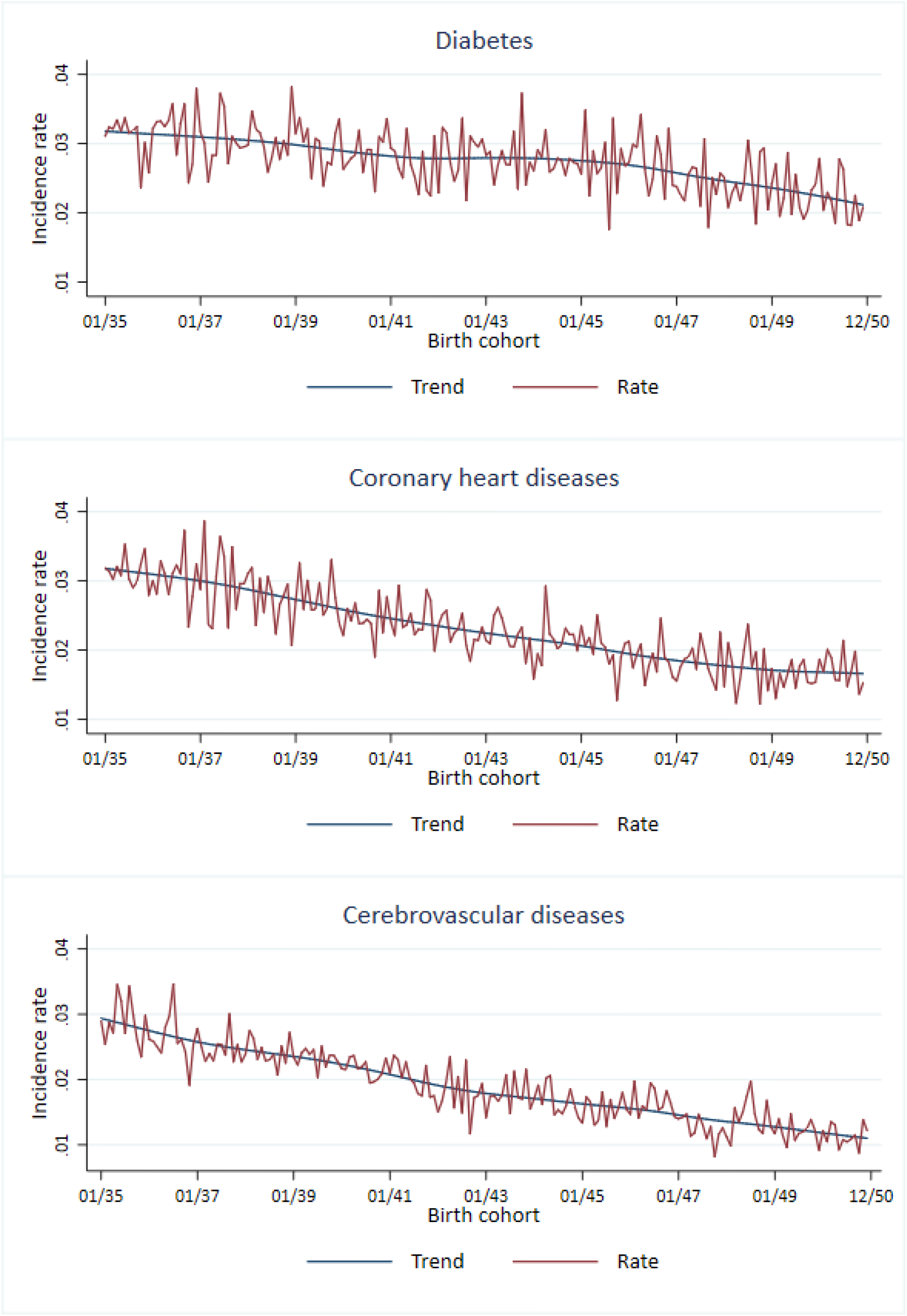
Rate and trend in diabetes, CHD, and CeVD incidence by birth cohort Data source: AOK 2004-2015

The recursive partitioning method resulted in stable nodes. For diabetes, the median of the first node is 83 (25^th^ percentile=71/75^th^ percentile=86) and all 1,000 replications split the data accordingly (NA=0). The second node is 34 (29/42,5) with only one replication not finding this node (NA=1). The third is 118 (108/130), and 6 replications do not find this node. Similar classification trees are derived for CHD (second node=76 (69/84), NA=0; second node=29 (23/40), NA=15; third node=117 (111/138), NA=35). For CeVD the results are less stable. While the first node is again found in all replications, the second and third nodes are only found by 88.7% and 89.4% respectively (second node=30 (24/36), NA=0; second node=66 (55/76), NA=113; third node=108 (100/124), NA=106). Overall, these three knots distinguish between cohorts born before the war, cohorts born during the first part and the second part, of the war, and cohorts born from the end of the war onwards. We use these to define four groups of birth cohorts depending on the disease of interest (diabetes: 1/35-10/37, 11/37-11/41, 12/41-10/44, 11/44-12/50; CHD: 1/35-5/37, 6/37-4/41, 10/44-12/50; CeVD: 1/35-6/37, 7/37-6/40, 7/40-12/43, 1/44-12/50) (Table 2).

**Table 2:**
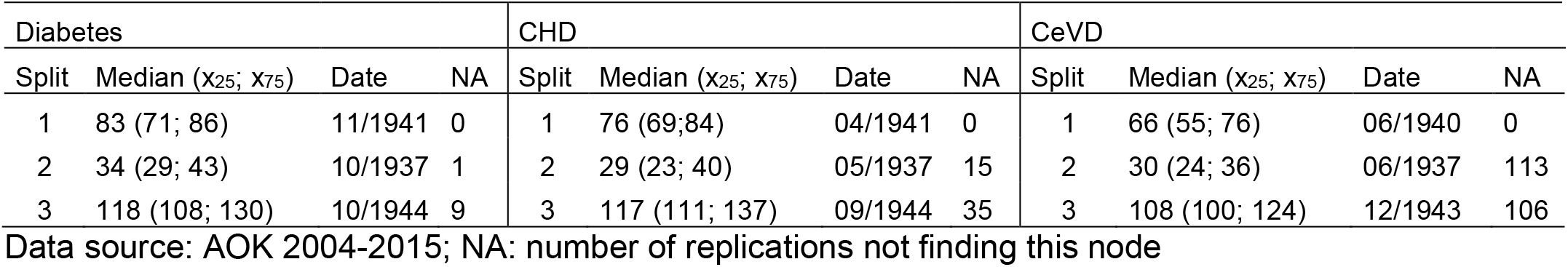
Median, 25^th^ and 75^th^ percentile from 1,000 bootstrap samples with 10,000 cases per sample

| Diabetes |  |  |  | CHD |  |  |  | CeVD |  |  |  |
| --- | --- | --- | --- | --- | --- | --- | --- | --- | --- | --- | --- |
| Split | Median (x<sub>25</sub>; x<sub>75</sub>) | Date | NA | Split | Median (x<sub>25</sub>; x<sub>75</sub>) | Date | NA | Split | Median (x<sub>25</sub>; x<sub>75</sub>) | Date | NA |
| 1 | 83 (71; 86) | 11/1941 | 0 | 1 | 76 (69;84) | 04/1941 | 0 | 1 | 66 (55; 76) | 06/1940 | 0 |
| 2 | 34 (29; 43) | 10/1937 | 1 | 2 | 29 (23; 40) | 05/1937 | 15 | 2 | 30 (24; 36) | 06/1937 | 113 |
| 3 | 118 (108; 130) | 10/1944 | 9 | 3 | 117 (111; 137) | 09/1944 | 35 | 3 | 108 (100; 124) | 12/1943 | 106 |

Data source: AOK 2004-2015; NA: number of replications not finding this node

We now turn to the Cox-regression models (Table 3). We found a general decline in cohort incidence for all three diseases, however, there was a discontinuation for diabetes among cohorts born in the second period of WWII (12/41-10/44), and deceleration of the decline for cohorts born from the end of WWII onwards for CeVD (11/44-12/50).

**Table 3:**
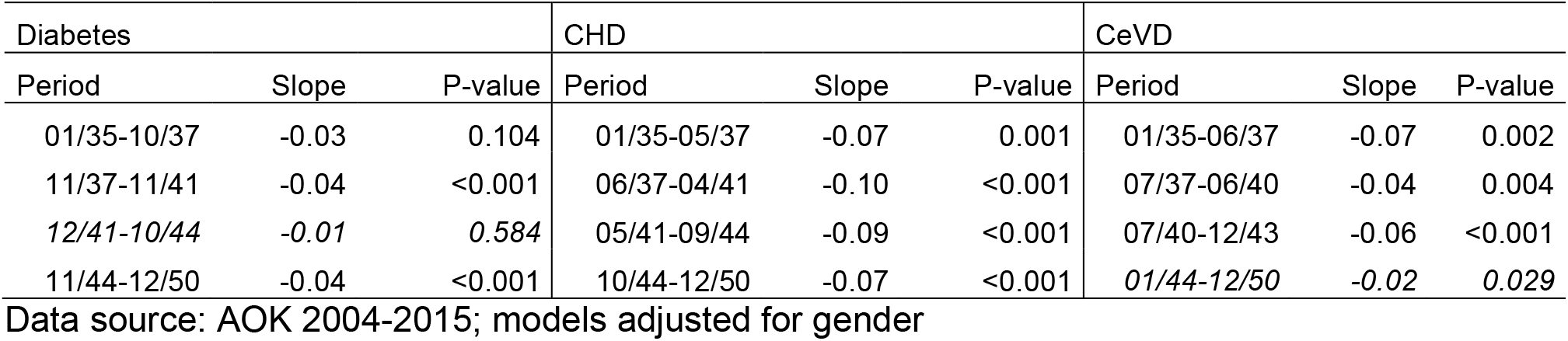
Parameter estimates of the yearly slopes of the linear splines from Cox Proportional Hazard Models

Regarding diabetes, there was a downward trend in the period 1/35-10/37 (yearly slope=-0.03, P=0.104), however, this was not statistically significant. The downward trend was significant among cohorts 11/37-11/41 (yearly slope=-0.04, P=0.001). There was a break in the period thereafter (12/41-10/44, yearly slope=-0.01, P=0.584). After this period, we found a downward trend again in 11/44-12/50 (yearly slope=-0.04, P<0.001).

Exploring the trend in CHD incidence, we found a negative yearly slope among all cohorts 1/35-5/37 (yearly slope=-0.07, P=0.001), 6/37-4/41 (yearly slope=-0.10, P<0.001), 5/41-9/44 (yearly slope=-0.09, P<0.001), and 10/44-12/50 (yearly slope=-0.07, P<0.001).

Similar to CHD, the trend in the incidence of CeVD was declining over all periods, by yearly slopes of −0.07 (P=0.002) among cohorts 1/35-6/37, −0.04 (P=0.004) among cohorts 7/37-6/40, −0.06 (P<0.001) among cohorts 7/40-12/43, and −0.02 (P=0.029) among cohorts 1/44-12/50. Calculating the marginal effect, we tested whether the deceleration of the decline in the last period differed significantly from the decline in the periods before the end of WWII, which it did (P=0.054).

We found similar patterns for each of the diseases in separate models for men and women (results not shown).

## Discussion

German claims data enabled us to explore the trend in the incidence of diabetes, CHD, and CeVD in birth cohorts 1935-1950 based on a study sample of about 75,000-90,000 cases, depending on the disease of interest. We identified cohorts born during the second part of WWII, which have not experienced a downward trend in the incidence of diabetes to the same extent as other cohorts before or after. We did not find significant results regarding the incidence of CHD and a significant deceleration in the decline of CeVD for cohorts born at the end of WWII and thereafter.

Two approaches for segmentation of birth cohorts regarding their risks of late-life disease outcomes are possible. One approach is to segment the cohorts by a-priori, historical defined criteria as applied by Stephan, et al. ^23^. In our data-driven approach we used statistical learning methods to define cut-points and to cluster cohorts with similar risk profiles. This proceeding may be better to consider the structure and characteristics of the data, and may lead to more precise predictions of cut-points. Consistent with our hypothesis, our results may suggest that conditions around birth and in the first years of life have long-term effects on late-life chronic and severe diseases.

In Germany, conditions during the second part of WWII had deteriorated in terms of housing, infrastructure, caloric intake, and nutrition; war-related stress had increased. Even if severe hunger periods only occurred after the end of WWII, this deterioration may be related to long-lasting deficiencies in health. Adverse conditions during gestation and in the first years of life have been shown to be associated with negative health outcomes later in life. Foetal programming^24^ by the social and biological environment may result in a thrifty phenotype^25^, subsequently leading to later life metabolic diseases such as stroke, diabetes, and hypertension, as well as other health measures^26-28^. Adverse socioeconomic conditions^29^ and a harsh family climate in childhood^30^ are known to influence levels of inflammation markers in adolescence and adulthood, possibly through changes in gene expressions. Early-life stress may thus engender a chronic activation of inflammatory pathways and increases the risk of diabetes, stroke, cardiovascular diseases, and the metabolic syndrome^31^. Early-life adversity is also associated with accelerated telomere length attrition as an indicator of cell aging^32^, which itself is associated with complications of type 2 diabetes^33^ or stroke^34^.

Regarding the risk of diabetes in our study, the incidence over cohorts 12/1941-10/1944 did not follow the declining trend as compared to birth cohorts before or after. These cohorts were affected by war-related adverse conditions during their first years of life, including gestation. They were also affected by the hunger crisis (from late 1944-48 in Germany) during their first years of life, but not during gestation (Figure 2). Further research may aim to analyse whether the effects are stronger at higher ages, which, however, would require a longer study period.

**Figure 2:**
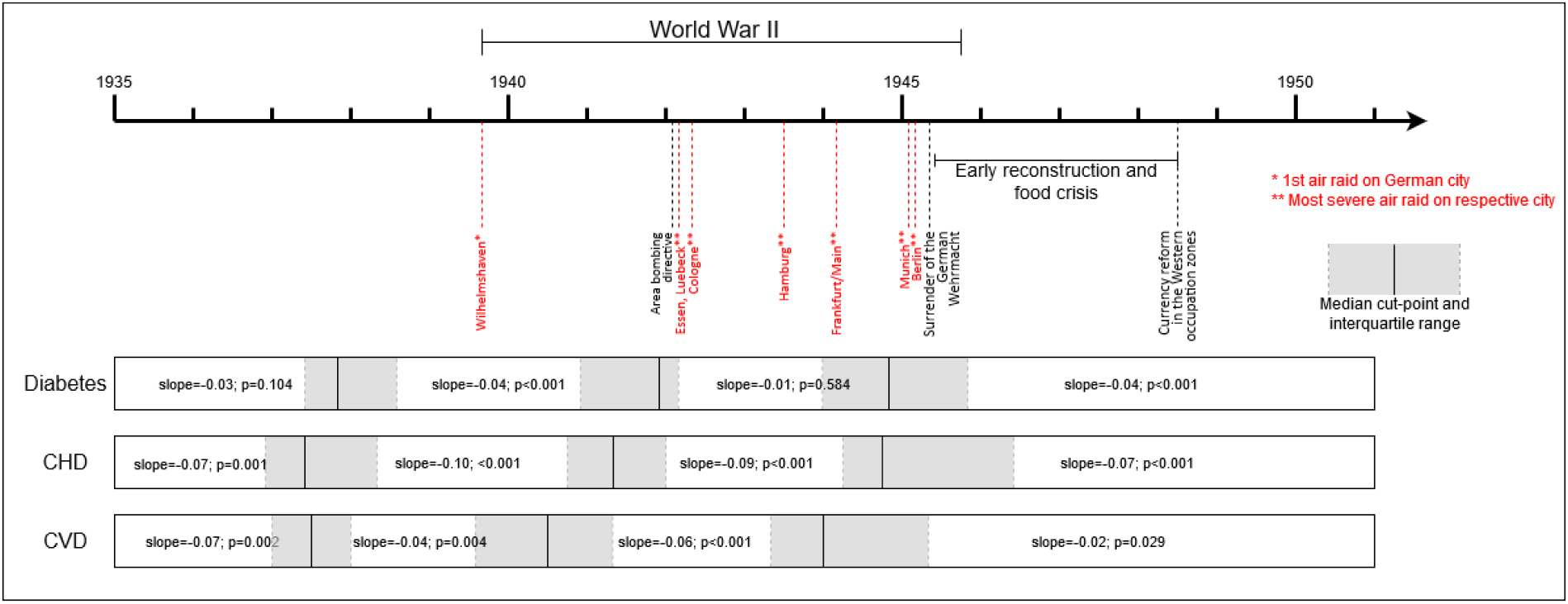
Cohorts according to diabetes, CHD, and CeVD risk profile from recursive partitioning in Germany between 1935-1950 Source: Own presentation

The lack of negative consequences for cohorts born during the hunger period may be associated with survival of the fittest, i.e. that more vulnerable offspring died before entering the relevant ages of later-life diseases due to the impact of the famine particularly on the foetus.

As a consequence, the frailty distribution of the population is altered because the remaining survivors are healthier.

One major drawback of claims data lies in their failure to provide detailed demographic and socioeconomic information. For example, we have no information on place of birth or nationality, hence we cannot differentiate between native- and foreign-born persons. Immigrants may have experienced different living conditions at birth and over their life-courses in their countries of origin. In contrast to Germany, not every country has experienced critical historical events such as WWII. However, the largest migrant groups in Germany stem from Italy, Poland, Greece, Romania, and Turkey^35^. All of these countries also experienced effects of WWII, closely associated with increased mortality, hunger, and stress^36-40^. Although this may lead to similar late-life effects on morbidity and mortality compared to the German population, it is possible that different timing of exposure may affect our results.

Our study could not confirm earlier results on the association between early-life effects on the risk of CHD. It remains unclear whether this is related to limitations of our study, such as potential selection effects, or to the composition of the sample as described above. The results may also be affected by missing information in our data. In addition to place of birth or nationality, the data do not provide lifestyle, socio-demographic, and medical information, such as intensity of former or current tobacco use, dietary habits, or body mass index, which could potentially affect the association between early-life conditions and later life health. People insured through the AOK have a lower overall socio-economic status compared to people insured in other public health insurance companies and privately insured persons, but this is more relevant among younger people^41,42^. Changes in the social status composition of the insured population are negligible in the time period studied here. The comparison of mortality patterns among the AOK and the German population indicates only marginal differences^43^.

Cohort–related selection effects into public health insurance may also have an effect, because people with private insurance tend to be healthier^44^. While we are not aware of cohort-specific changes in entry rules to private insurance, health selection may lead to an underestimation of the effect observed in our study. If certain WWII cohorts had generally worse health, then they would have been barred from entering private health insurance, which in our data would make them less selected for positive health than previous or following cohorts.

Finally, diagnoses in medical claims data are neither specific nor standardized, and a claims-based definition of diagnoses is not the same as prospective clinical assessment. We tried to cope with the latter by implementing an internal validation procedure (see Data and Methods).

The strength of our study lies in the data driven approach to identify risk periods associated with WWII, and the nationwide population-based dataset with a large sample size. The analysis of health claims data instead of population-based surveys avoids potential biases, such as response behaviour or self-selection, selection by the health care provider or the study design. Of note is that community-dwelling and people living in nursing homes are included in this study, as particularly the latter are usually missing in surveys.

Our study suggests that the second period of WWII interfered with the cohort decline in the risk of diabetes, and that the hunger period from the end of WWII onwards may have decelerated the decline in CeVD. We explored diseases that are closely associated with mortality, disability, and poor quality of life, resulting in high costs for the health care system. This is also of particular importance because our focus was on diseases that are known risk factors for or comorbidities associated with cognitive decline and dementia^45-48^. Thus, negative outcomes in terms of the metabolic and vascular diseases might subsequently lead to an increased risk of dementia. Especially in the context of ageing societies, our results support the crucial role of critical events in the first years of life and support the importance of health, family, and social policies directed towards disadvantaged individuals, in order to avoid negative long-term consequences on the health status.

## Data Availability

The scientific research institute of the AOK (WIdO) has strict rules regarding data sharing because of the fact that health claims data are a sensible data source and have ethical restrictions imposed due to concerns regarding privacy. Anonymized data are available to all interested researchers upon request. Interested individuals or an institution who wish to request access to the health claims data of the AOK, please contact the WIdO (webpage: http://www.wido.de/, mail).

## Acknowledgements

We are grateful to Juergen-Bernhard Adler, Andreas Kloess, and Christian Guenster from the Scientific Research Institute of the AOK (Wissenschaftliches Institut der AOK - WIdO) for providing the data. The authors are also thankful to Renée Lüskow for her English editing service. We thank the participants at a workshop on Public Health Economics in Groningen (2019) for a fruitful discussion.

